# Reduced sensitivity of SARS-CoV-2 Omicron variant to booster-enhanced neutralization

**DOI:** 10.1101/2021.12.17.21267961

**Authors:** Xiaoqi Yu, Dong Wei, Wenxin Xu, Yulong Li, Xinxin Li, Xinxin Zhang, Jieming Qu, Zhitao Yang, Erzhen Chen

## Abstract

The Omicron Variant of Concern (B.1.1.529) has spread internationally and is raising serious concerns about reduced vaccine efficacy and the increased risk of reinfection. We assessed the serum neutralizing activity using a pseudovirus-based neutralization assay in 292 healthcare workers who had received a homologous booster dose of BBIBP-CorV vaccine, 8 to 9 months after completing the priming two-dose vaccination schedule, to investigate whether the newly identified Omicron variant can escape serum antibody neutralization induced by the booster vaccination. The booster dose of BBIBP-CorV rapidly induced a significantly high level of humoral immune response, and the neutralization geometric mean titer (GMT) against the wild-type strain on day 28 after the booster dose was 294.85 (252.99-343.65), 6.1 times higher than the level on day 28 after the second dose. The neutralization against the Omicron variant was also improved by the booster vaccination, although the GMT showed an approximately 20.1-fold reduction to 14.66 (12.30-17.48) when compared with the wild-type strain. This study demonstrated that a booster dose of BBIBP-CorV led to a significant rebound in neutralizing immune response against SARS-CoV-2, while the Omicron variant showed partial resistance to neutralizing antibodies induced by the booster vaccination.

---

As of December, 2021, more than 272 million people have been infected with SARS-CoV-2. Multiple types of vaccines have been used to build herd immunity for the pandemic, however, decreased protective effect has been reported, and neutralizing antibody titers induced by the two doses of vaccination decline to near or below the seropositive threshold after 6 months^1^, indicating relatively short-duration protection provided by the current COVID-19 vaccines. Additionally, with the unprecedented transmission of SARS-CoV-2, several more contagious Variants of Concern (VOCs) have emerged. Most recently, the B.1.1.529 variant Omicron, which was identified in November 2021, has spread internationally. The Omicron variant is the fifth VOC designated by the World Health Organization, primarily due to numerous mutations in the spike glycoprotein, especially in the receptor-binding domain and N-terminal domain. As the Omicron variant is the most divergent variant so far, it may lead to escape from immunity induced by the existing COVID-19 vaccines, and cause a large number of breakthrough infections^2^.

Waning immunity and viral diversification both create the potential need for further booster vaccination, therefore, we administered a homologous booster dose of the BBIBP-CorV vaccine, 8 to 9 months after completing the priming two-dose vaccination schedule, to eligible healthcare workers in Shanghai Ruijin Hospital to investigate whether the newly identified Omicron variant can escape serum antibody neutralization induced by the booster vaccination. Serum specimens were obtained 28 days after the second dose, before and 28 days after the booster dose. We determined the serum neutralizing activity using a pseudovirus-based neutralization assay, and SARS-CoV-2 specific antibody level, which is thought to be a good surrogate for neutralizing antibodies, were also assessed using a chemiluminescence immunoassay. The details of the methods were described in the Supplementary Information.

A total of 292 participants were included in this study, of whom 72 were male and 220 were female, with a median age of 39.00 years (interquartile range [IQR] 32.00-46.00) years. The baseline immune responses at 8 to 9 months after the priming vaccination with two doses were weak. Specific antibodies against SARS-CoV-2 could still be detected in 229 (78.42%) of 292 participants, but the median antibody level dropped from 31.98 (10.36-73.66) on day 28 after the two-dose vaccination to 3.63 (1.16-9.93) (Fig 1a). Moreover, only 53 (18.15%) of 292 participants had quantifiable neutralizing antibodies over the period of 8-9 months, and the geometric mean titer (GMT) declined rapidly to below the lower limit of detection (Fig. 1b).

**Figure 1:**
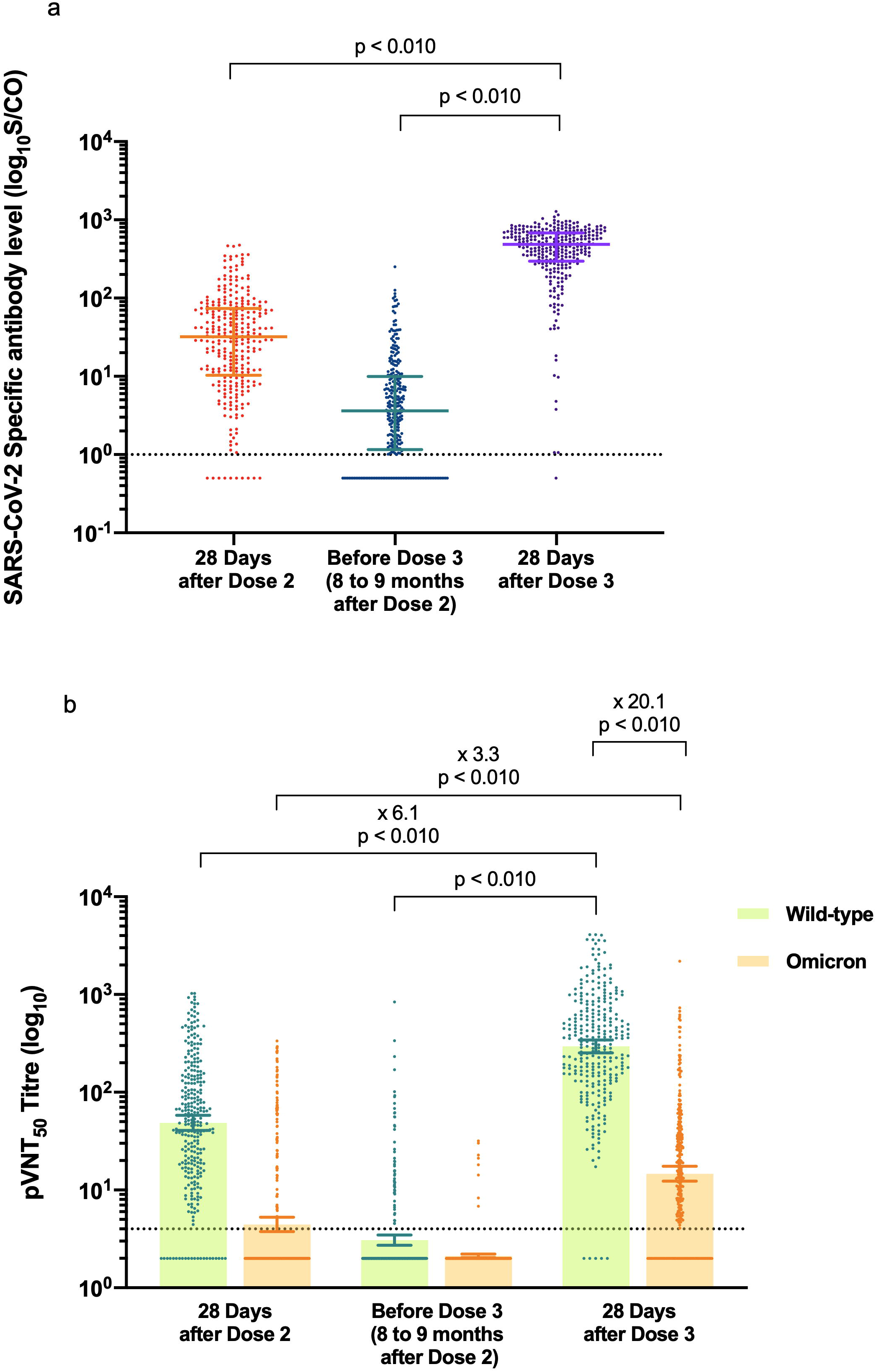
Anti-SARS-CoV-2 specific antibody and neutralizing antibody responses in serum samples of the 292 participants. **a**, The specific antibody levels against SARS-CoV-2 at different time points are shown. The horizontal dashed line represents the lower limit of detection (LLD) of 1. Results below the LLD were set to 0.5 times the LLD. Data points shown on the bar graph represent individual titers. Error bars indicate median and interquartile range (IQR). **b**, Results of 50% pseudovirus neutralization titer (pVNT50) against the wild-type strain and the Omicron variant at the time points are shown. The horizontal dashed line represents the lower limit of detection (LLD) of 4. Results below the LLD were set to 0.5 times the LLD. Data points shown on the bar graph represent individual titers. Error bars represent the geometric mean with the 95% confidence interval (95% CI). Fold-changes in geometric mean titer are shown above. p values were calculated using the Wilcoxon matched-pairs signed-rank test.

A significantly enhanced antibody response was observed on day 28 after the booster dose. Specific antibodies against SARS-CoV-2 against SARS-CoV-2 were detected in 291 (99.66%) of 292 participants, with a median antibody level of 486.66 (296.16-681.91), which was markedly higher than the baseline antibody level and the level on day 28 after the second dose (Fig 1a). The seroconversion rate of neutralizing antibodies against the wild-type strain was 98.29% (287/292), and the GMT increased to 294.85 (95% CI 252.99-343.65), more than 6.1 times of the level on day 28 after the second dose (Fig. 1b). The GMT on day 28 after the second dose was 48.65 (40.67-58.19), with 270 (92.47%) of 292 individuals showing detectable neutralization.

On day 28 after the second dose, 75 (25.68%) of 292 vaccinated individuals displayed detectable serum neutralizing antibodies against Omicron variant, resulting in a GMT of 4.45(3.75-5.28). Over the period of 8-9 months, only eight out of 292 vaccinated individuals displayed quantifiable neutralizing antibodies against Omicron variant before the booster dose. Notably, on day 28 after the booster dose, 228 (78.08%) participants had neutralizing activity against the Omicron variant, and the booster dose resulted in approximate 3.3-fold improvement in neutralization against the Omicron variant compared with the second vaccination, although the GMT showed a 20.1-fold reduction to 14.66 (12.30-17.48) relative to the wild-type strain (Fig. 1b), demonstrating partial resistance of Omicron variant to neutralizing antibodies elicited by vaccination. Additionally, Sex and age were not factors that associated with the induction of neutralizing antibody and neutralizing titers against SARS-CoV-2 and the Omicron variant after the booster dose.

Recent study has reported that a third dose of BNT162b2 mRNA vaccine is effective in preventing severe COVID-19 outcomes^3^. The booster dose of either inactivated vaccine or recombinant protein subunit vaccine can quickly recover the neutralizing immune response to SARS-CoV-2^4^, and elicit neutralizing antibodies against VOCs^5^. This study aimed to determine whether a homologous booster dose of inactivated vaccine can effectively activate specific immune responses to SARS-CoV-2, especially enhancing the neutralizing activity against the newly-emerged Omicron variant. The data revealed that approximately 8 to 9 months after the priming with two doses of inactivated vaccine, the neutralizing activity declined rapidly and could hardly be detected, supporting the need for a third dose to extend the duration of the humoral immune response against the emerging variants. As expected, a third dose following the priming with two doses of inactivated vaccine significantly recalled and enhanced antibody responses, indicating that the priming vaccination could induce efficient memory humoral immune responses. The neutralization GMT against the wild-type strain on day 28 after the third dose was 6.1 times higher than the GMT on day 28 after the second dose, but the persistence of the enhanced immunity against SARS-CoV-2 and its variants induced by the booster vaccination remains to be evaluated.

It has been suggested that achieving a higher neutralizing antibody titer with a booster dose is desirable to increase the breadth of neutralization^6^. However, based on the current data, sera from vaccinated and convalescent individuals neutralized the Omicron variant to a much lesser extent than any other VOC^7^. The neutralization capacity of vaccine-elicited sera against the Omicron variant was reduced at least 10-fold^8^, and the 8.4-fold reduction in the neutralization of convalescent sera suggested that the Omicron variant may escape from immune protection elicited by previous SARS-CoV-2 infection and by vaccination with existing COVID-19 vaccines^9^. In our findings, the booster vaccination significantly improved the humoral immune response against the Omicron variant, which might be associated with the higher magnitude of wild-type neutralization, although the neutralizing activity was much less effective against the Omicron variant, with an approximately 20.1-fold reduction in neutralization titers relative to the wild-type strain. Since neutralization is only a part of the humoral immune response, and neutralizing activity does not reflect all potentially protective immune responses, real-world studies regarding the protection efficacy of the booster vaccination against the Omicron variant are still needed.

In conclusion, a booster dose of BBIBP-CorV led to a significant rebound in neutralizing immune response against SARS-CoV-2, while the Omicron variant showed extensive but incomplete escape from booster-enhanced neutralization. In current situation with the Omicron variant causing a rapidly increased number of infections, the data presented here contribute evidence toward establishing a booster vaccination strategy against COVID-19.

## Data availability

The data that support the findings of this study are available from the corresponding authors on reasonable request.

## Acknowledgments

This study was funded by the Shanghai Key Laboratory of Emergency Prevention, Diagnosis and Treatment of Respiratory Infectious Diseases (20dz2261100), and a grant from the Science and Technology Commission Shanghai Municipality (No. 20JC1410200). The investigators express their gratitude to Prof. Guang Ning for his great support. We thank all the participants involved in this study.

## Author contributions

XZ, JQ, ZY, and EC conceived and designed the study. XY and WX were responsible for collecting and summarizing the clinical data. DW, WX, YL, and XL performed the experiments. XY and DW carried out the analysis. XY and XZ drafted the manuscript. All authors reviewed and approved the final version.

