## Supplementary Methods for "Reduced sensitivity of SARS-CoV-2 Omicron variant to booster-enhanced neutralization"

**Supplementary Information, Materials and methods.**

**Participants and blood samples**

Eligible participants for the third booster dose were healthcare workers in Shanghai Ruijin Hospital aged 18-59 years, who have received priming vaccination by two doses, 21 days apart of inactivated SARS-CoV-2 vaccine (BBIBP-CorV, Sinopharm) for at least 8 months. Blood samples were taken from 292 participants for serology tests before and 28 days after the third booster dose. The protocol and informed consent were approved by the Ethics Committee of Shanghai Ruijin Hospital (RJHKY2021-321) in accordance with the Declaration of Helsinki and Good Clinical Practice. Written informed consent from all participants was obtained.

**SARS-CoV-2 specific antibody response assessment**

The specific antibodies against SARS-CoV-2 were measured using a chemiluminescence kit manufactured by Wantai BioPharm (China). The antibody levels were expressed by the chemiluminescence signal according to the manufacturer’s instructions.

**Pseudovirus based neutralization assay**

A pseudovirus-based neutralization assay was performed as previously described^18^ with minor modifications. In brief, the lentivirus-based SARS-CoV-2 pseudoviruses expressing a luciferase reporter gene and bearing the spike protein from the wild-type strain (Wuhan-Hu-1) and the Omicron variant (lineage B.1.1.529) tested in this study were manufactured by Vazyme Biotech Co., Ltd (China). Vaccine-elicited sera were inactivated at 56℃ for 30 mins before assess the neutralization geometric mean titers (GMTs). Six serial dilutions of heat-inactivated sera (in a four-fold step-wise manner, initially 1:4 diluted) were incubated with 250 TCID_50_ SARS-CoV-2 pseudoviruses per well for 1 hour, together with the virus control and cell control wells, before seeding 20,000 HEK293T-ACE2 cells per well in 96-well plates. Following 48 hours of incubation in a 5% CO_2_ environment at 37℃, the supernatant was removed, and the luminescence were measured using Luciferase Assay System (Promega Biotech Co., Ltd) according to the manufacturer’s instructions. The 50% inhibitory concentration (IC_50_) was defined as the serum dilution at which the relative light units (RLUs) were reduced by 50% compared with the virus control wells after subtraction of background RLUs in the cell control wells with only cells. The IC_50_ values were calculated by generating a three-parameter non-linear regression curve fit in GraphPad Prism 8.4.0. Neutralizing antibody potency < 1:4 was considered negative. The neutralizing titer for each sample was measured twice in two independent experiments.

**Statistical analysis**

Continuous variables that were not normally distributed were presented as median (interquartile range [IQR]). Categorical variables were described as count (%). The statistical method used for analysis of antibody titers is the geometric mean (GMTs) and the corresponding 95% confidence interval (95% CI). The values were compared by the Wilcoxon matched-pairs signed-rank test. Graphs were plotted using GraphPad Prism 8.4.0. Statistical analyses were done using SPSS 24.0. A two-sided *P* value of less than 0.050 was considered statistically significant.
